# Sleep moderates the effects of exercise on cognition in chronic stroke: secondary analysis of a randomized trial

**DOI:** 10.1101/2024.01.16.24301392

**Authors:** Ryan S. Falck, Ryan G. Stein, Jennifer C. Davis, Janice J. Eng, Laura E. Middleton, Peter A. Hall, Teresa Liu-Ambrose

## Abstract

**Background:** Exercise (EX) or cognitive and social enrichment (ENRICH) are two promising strategies for promoting cognitive function post-stroke. Sleep may moderate the effects of these interventions on cogntion, whereby intervention effects may be more robust among individuals suffering from poor sleep. We examined whether sleep moderates the effects of EX or ENRICH on cognitive function in adults with chronic stroke.

**Methods:** Secondary analysis of a three-arm parallel, single-blinded, randomized clinical trial among community-dwelling adults aged 55+ years with chronic stroke (i.e., ≥12 months since stroke). Participants were randomized to 2x/week EX, ENRICH, or balance and tone control (BAT). At baseline, device-measured sleep duration and efficiency were measured using MotionWatch8 wrist-worn actigraphy; self-reported quality was measured by Pittsburgh Sleep Quality Index (PSQI). Participants were categorized at baseline as having good or poor: 1) device-measured duration; 2) device-measured efficiency; or 3) self-reported quality. The primary cognitive outcome was Alzheimer’s Disease Assessment Scale Plus (ADAS-Cog-Plus) measured at baseline, 6 months (end of intervention), and 12 months (6-month follow-up). Linear mixed models examined if baseline sleep categorizations (i.e., good/poor) moderated the effects of EX or ENRICH on ADAS-Cog-Plus.

**Results:** We enrolled 120 participants in the trial (EX=34; ENRICH=34; BAT=52). Baseline sleep categorization did not moderate the effect of ENRICH on ADAS-Cog-Plus; however, it moderated the effect of EX. EX participants with poor baseline sleep efficiency (estimated mean difference: −0.48; 95% CI:[−0.85, −0.10]; *p*=0.010) or self-reported sleep quality (estimated mean difference: −0.38; 95% CI:[−0.70, −0.07]; *p=*0.014) had significantly better ADAS-Cog-Plus performance at 6 months compared with BAT participants with poor sleep. There was no effect of EX on ADAS-Cog-Plus for participants with good baseline sleep.

**Conclusion:** The effects of EX on cognitive function in adults with chronic stroke is moderated by sleep, whereby poor sleepers benefit more.

## Introduction

A stroke occurs every 40 seconds.^1^ A stroke doubles one’s risk for dementia,^2^ and stroke-related cognitive deficits are associated with reduced functional independence, institutionalization, reduced quality of life, and early death.^3^ Stroke survivors need targeted interventions to promote cognitive function and prevent dementia. Two promising strategies for promoting cognitive health in stroke survivors are: 1) exercise training; and 2) cognitive and social enrichment.^4^

Exercise training (EX) is defined as planned or structured physical activity with the intent of increasing or maintaining physical fitness.^5^ While the precise prescription of EX (i.e., type, intensity, or volume of exercise) for enhancing cognitive function is still uncertain, evidence from randomized clinical trials (RCTs) suggests that moderate-or-higher intensity EX improves cognitive function and promotes better brain health among older adults.^6^ ^7^

Cognitive and social enrichment (ENRICH) is broadly an intervention designed to increase cognitive and social activity by provision of a stimulating environment.^8^ The premise of this strategy is that engaging in ENRICH activities can stimulate higher-order cognitive processes (e.g., memory and executive function), and thus promote better cognitive function. The use of ENRICH to promote cognitive health in stroke survivors is supported by evidence from both animal and human studies.^8^ ^9^

In our previously published primary study,^4^ we showed multi-component moderate-intensity EX induced clinically important improvements in cognitive function in adults with chronic stroke; in contrast, ENRICH did not. However, a large degree of variation exists in the efficacy of lifestyle interventions. For example, a meta-analysis determined that effect size estimates for the impact of EX on cognitive function varied between a Hedge’s *g* of −1.89 and 3.41. To maximize the benefits of lifestyle interventions for cognitive health, we need to identify key moderators which can maximize efficacy.

Sleep is a potential moderator of the effects of lifestyle interventions on cognitive function. Poor sleep is common following stroke.^10^ Approximately 40% of stroke survivors have diagnosed sleep disorders such as excessive daytime sleepiness, obstructive sleep apnea (OSA), and insomnia.^11^ ^12^ Poor sleep quality among stroke survivors increases the risk of recurrent stroke by 3-fold and the risk of early death by 76%.^13^ Of relevance, adults with chronic stroke (i.e., ≥12 months since stroke sequelae) and poor sleep also experience larger deficits in cognitive performance compared with their counterparts without sleep problems.^14^

In this secondary analysis, we examined whether baseline sleep quantity or quality moderates the effects of EX or ENRICH on cognitive function in adults with chronic stroke. We hypothesized that baseline sleep quantity and quality would moderate the effects of EX on cognitive function. Specifically, EX would improve cognitive function among those with poor sleep but not among those with good sleep. Given that there was no effect of ENRICH on cognitive function in the primary study,^4^ we hypothesized that baseline sleep would not moderate the effects of this intervention.

## Methods

### Study Design

This was a secondary analysis of a three-arm parallel, single-blinded, 6-month RCT with a 6-month follow-up in a research centre (Vancouver, British Columbia, Canada) to examine the effects of EX or ENRICH activities on cognitive function in community-dwelling adults with chronic stroke.^4^ Participants were measured at baseline, at the end of the 6-month intervention, and at 6-month follow-up (Figure 1). Ethical approval was provided by University of British Columbia’s Clinical Research Ethics Board (H13-00715) and Vancouver Coastal Health Research Institute (V13-00715). The trial protocol and the primary study results are published (ClinicalTrials.gov identifier: NCT01916486).^4^ ^15^

**Figure 1.**
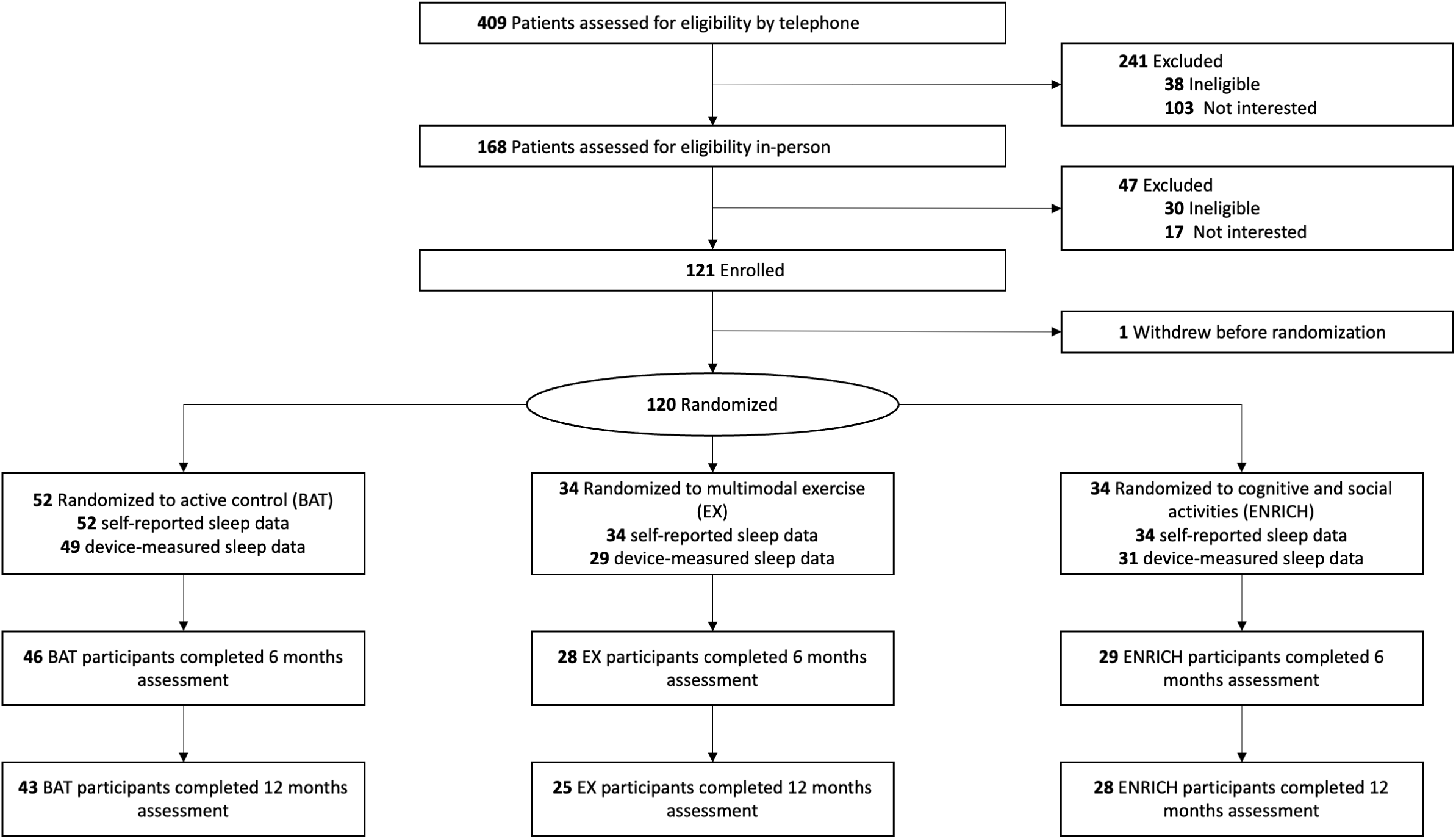
CONSORT Diagram.

### Recruitment

Participants were recruited from the community as well as from stroke clinics. Enrollment and randomization occurred from June 6, 2014 to February 26, 2019.^4^

### Inclusion and Exclusion Criteria

We included community-dwelling adults who had an ischemic or hemorrhagic stroke. Additional inclusion criteria were: 1) aged ≥ 55 years; 2) a history of a stroke ≥ 12 months prior to study enrollment; 3) a Mini-Mental State Examination (MMSE)^16^ score of ≥ 20/30 at screening, including a perfect score on the 3-step command to ensure intact comprehension and ability to follow instructions; 4) English speaking; 5) not expected to start or were on a stable fixed dose of cognitive medications during the study period; 6) able to walk six metres with rest intervals with or without assistive devices; and 7) not currently participating in any regular therapy or progressive exercise. Exclusion criteria were neurodegenerative disease, dementia, at high risk for cardiac complications during exercise, taking medications that may negatively affect cognitive function, or aphasia as judged by an inability to communicate by phone. For our analyses, we included all participants with available sleep data at baseline. All participants provided written informed consent.

### Randomization and Blinding

Participants were stratified by stroke status (one versus ≥ two stroke events) and randomly allocated to: 1) multi-component EX; 2) ENRICH; or 3) an active control group consisting of stretching and toning exercises (BAT) with an allocation ratio of 2:2:3 (EX:ENRICH:BAT, respectively) using permuted blocks within each stratum. The allocation ratio accounts for the two planned contrasts using the Dunnett Test.^17^ Allocation was concealed.

Assessors were blinded to participants’ allocation and participants were asked to refrain from discussing their study involvement or experience during assessments with assessors. Participants and those who delivered the interventions were not blinded.

### Sample Size Calculation

Our sample size was powered to evaluate the treatment effect between-groups on the Alzheimer’s Disease Assessment Scale Plus (ADAS-Cog-Plus) at the end of the six month intervention.^15^ We assumed a standardized effect size of 0.6 of exercise on cognitive function based on our prior work using the 11-item ADAS-Cog.^18^ Assuming an alpha of 0.05, 39 participants per group (i.e., total sample of 117) would provide a power greater than 0.80. We then assumed a standardized effect size of 0.7 for the ADAS-Cog-Plus, given it has greater sensitivity to changes in cognition compared with either the 11- or 13-item ADAS-Cog.^19^ After accounting for 15% attrition and 2:2:3 allocation, 34 participants were randomized to EX, 34 to ENRICH, and 52 to BAT, for a total sample of 120.

### Interventions

Each treatment arm included twice-weekly supervised classes of 60 minutes over six months and have been described previously.^15^ All instructors were trained by the research team over a three-hour session and delivered the interventions based on written protocols.

EX was a multi-component exercise intervention based on the Fitness and Mobility Exercise program (www.fameexercise.com).^20^ It included strength training, aerobic, agility, and balance exercises. Intensity of aerobic training was monitored by the Borg Rating of Perceived Exertion (RPE) and heart rate monitors.

ENRICH included computerized cognitive training,^21^ other activities which used apps, and others which were based on improvisation and mental activities from the Perk activities program.^22^ It was designed based on a prior pilot study^23^ and current evidence.^24^

BAT consisted of stretches, deep breathing and relaxation exercises, general posture education, grip strength and dexterity exercises, and light isometric toning exercises.^25^ Once a month, an education seminar replaced other activities.

### Measures

We report measures acquired at baseline, six months (i.e., end of intervention), and 12 months (i.e., 6-month follow-up). All assessments were conducted by blinded assessors.

The Functional Comorbidity Index^26^ measured the number of comorbid conditions. Global cognitive function was assessed by MMSE.^16^ The Center for Epidemiologic Studies Depression Scale assessed for depressive symptoms.^27^ Motor function of the upper and lower extremities was assessed by the Fugl-Meyer Assessment Motor scale.^28^

#### Cognitive Function

Our primary outcome was the ADAS-Cog-Plus.^19^ Lower ADAS-Cog-Plus scores represent better cognitive performance. We included the 13-item ADAS-Cog^29^ (ADAS-Cog-13) as a secondary outcome, wherein a change of 3.0 points is a minimally important difference.^30^

#### Device-Measured Sleep Quantity and Quality

Device-measured sleep quantity and quality were indexed using the MotionWatch8© (MW8), a uni-axial, wrist-worn accelerometer with evidence of validity and reliability.^33,34^ We used 60 second epochs which is consistent with current guidelines for estimating sleep.^37^

At baseline, participants were fitted with the MW8 and provided detailed information on its features (i.e., the light sensor, event marker button, and status indicator). Participants were instructed to press the event marker button each night when they started trying to sleep; and again each morning when they finished trying to sleep. The established protocol for wrist-worn actigraphy suggests participants wear the MW8 on the non-dominant wrist for a period of 14 days^38^; however, this protocol was modified for older adults with stroke such that if the non-dominant side was the stroke-affected side, then we placed the MW8 on the dominant wrist.

Participants were also given the 9-item Consensus Sleep Diary (CSD) and asked to complete it each morning upon waking.^39^ The responses from the CSD were used to confirm sleep windows identified by participants.^40^ In cases where the event marker and CSD entry disagreed for the start time of the sleep window, we used activity cessation and light sensor data from the MW8 to determine “lights out.” Similarly, when the event marker and CSD entry disagreed for the end of the sleep window, we used activity onset and “lights on” to determine the end of the sleep window. If responses from the CSD entry disagreed with the event markers entered by participants as the start of the day (i.e., finished trying to sleep and awake and out of bed), we used activity onset and light sensor data to determine the start of the day. Similarly, when the event marker and CSD entry disagreed for the end of day (i.e., time spent trying to sleep), we used activity cessation and light sensor data to determine the end of the day.

Details of our data reduction procedure have been published.^41^ Briefly, data were analyzed using MotionWare 1.0.27 (cam*n*tech). Data prior to recorded wake-time on the first full day of recording were manually removed in order to only investigate full 24-hour recordings of activity. Each day of activity consisted of when the participant self-reported being awake and out of bed. Participant self-report was confirmed via event marker time stamps from MW8. The MotionWare software was then used to estimate sleep duration (i.e., total time spent sleeping) and sleep efficiency (i.e., time asleep expressed as a percentage of time in bed).

#### Self-reported Sleep Quality

We measured self-reported sleep quality using the Pittsburgh Sleep Quality Index (PSQI).^30^ The questionnaire surveys sleep quality spanning the previous month and has good evidence of validity and reliability.^30^

### Statistical Analyses

We performed all statistical analyses in R version 4.1.2 using the *psych, lme4,* and *lsmeans* packages. Our statistical code and output are available on <u>GitHub</u>. All models followed the intention-to-treat principle.

We classified participants as having good or poor: 1) device-measured sleep duration; 2) device-measured sleep efficiency; or 3) self-reported sleep quality at baseline. We indexed participants as having good baseline sleep duration as an average sleep duration 420-490 minutes/night at baseline ^31^; poor baseline sleep duration was categorized as anything outside of this range. Good baseline sleep efficiency was indexed as an average nightly efficiency ≥ 85%,^32^ while good baseline self-reported sleep quality was categorized as a PSQI score ≤ 5.^33^ Participants who failed to meet either the sleep efficiency or self-reported sleep quality criterion were classified as having poor efficiency or self-reported quality, respectively. Each of these criteria were chosen based on current guidelines for healthy sleep or epidemiological methods for classifying good versus poor sleepers.^31–33^

We then examined if treatment effects for our primary outcome, ADAS-Cog-Plus, were moderated by baseline device-measured sleep duration or efficiency or self-reported quality categorization (good/poor) using linear mixed models with restricted maximum likelihood estimation. The model included random intercepts, and fixed effects of time at baseline, end of the intervention, and 6-months follow-up, group assignment (i.e., EX, ENRICH, BAT), and their interaction. We conducted three separate models to address each sleep metric (i.e., device-measured sleep duration or sleep efficiency, or self-reported sleep quality); each model included baseline sleep categorization (i.e., good vs. poor) as a fixed effect, as well as its interaction with time and group assignment (i.e., sleep categorization x time x group). Baseline ADAS-Cog-Plus, MMSE score, and Fugl-Meyer motor score were included as fixed-effect covariates. Unequal variance was allowed across time, group, and sleep categorization. Estimated marginal means were calculated for each treatment group at baseline, end of the intervention, and 6-month follow-up based on sleep categorization.

We then performed planned contrasts using the Dunnett Test,^17^ a multiple comparison procedure, to assess differences in ADAS-Cog-Plus at end of the intervention and six-month follow-up between: 1) EX vs. BAT; and 2) ENRICH vs. BAT. The overall alpha was set at 0.05. Each contrast was estimated separately for good and poor baseline sleep categorizations. In the event of significant effects, we conducted post-hoc contrasts comparing the estimated between-group differences in ADAS-Cog-Plus for participants categorized as having good versus poor baseline sleep (i.e., Good Sleep – Poor Sleep). This was done in order to determine if baseline sleep categorization significantly moderated the effect of a given intervention.

Linear mixed models with restricted maximum likelihood estimation were also conducted on our secondary outcome of ADAS-Cog-13. Baseline value of outcome, MMSE score, and Fugl-Meyer motor score were included as fixed-effect covariates. Estimated marginal means were then calculated for each intervention group at baseline, end of the intervention, and 6-month follow-up based on sleep categorization. Planned contrasts using the Dunnett Test were then used to assess differences in outcome at the end of the intervention and 6-month follow-up, stratified by baseline sleep categorization. For significant effects, we then conducted post-hoc contrasts comparing the estimated between-group differences in ADAS-Cog-13 based on baseline sleep categorization (i.e., Good Sleep – Poor Sleep). Given the exploratory nature of our analysis, we did not control for multiple comparisons.

## Results

Figure 1 illustrates our CONSORT diagram. One-hundred and twenty participants were enrolled and randomized from June 6, 2014 to February 26, 2019. The final measurements were made on March 3, 2020 and were not impacted by the COVID-19 pandemic. Of the 120 participants randomized to this study, 109 participants (EX=29; ENRICH=31; BAT=49) completed 7 days observation of device-measured sleep at baseline. All 120 participants (EX=34; ENRICH=34; BAT=52) completed the PSQI at baseline. The attrition rate was 14% at the end of the 6-month intervention, and 20% at the end of the 6-month follow-up.

The mean baseline age of participants was 71 years (SD=9) and 62% were male (Table 1). Mean baseline ADAS-Cog-Plus score of 0.22 (SD=0.80), indicating that participants had cognitive impairment.^19^ The mean baseline Fugl-Meyer Assessment Motor score of 81.21/100 (SD=23.85) indicates moderate to mild motor impairment.^34^

**Table 1.** Participant Characteristics.

|  | EX<br>(N= 34) | ENRICH<br>(N= 34) | BAT<br>(N= 52) |
| --- | --- | --- | --- |
| Age | 70.65 (9.14) | 71.29 (9.25) | 70.44 (7.81) |
| Males n, % | 21, 61.8% | 23, 67.6% | 30, 57.7% |
| Body Mass Index (kg/m<sup>2</sup>) | 27.37 (3.69) | 27.32 (4.45) | 27.99 (5.05) |
| Level of Education n, % |  |  |  |
| *High School or Less* | 6, 17.6% | 5, 14.7% | 13, 26.5% |
| *Trade School or Some University* | 10, 29.4% | 8, 23.5% | 18, 36.7% |
| *University Degree or Higher* | 18, 52.9% | 21, 61.8% | 21, 40.4% |
| Number of Strokes | 1.09 (0.29) | 1.15 (0.50) | 1.29 (0.72) |
| Type of Stroke |  |  |  |
| *Hemorrhagic* | 8, 23.5% | 10, 29.4% | 15, 28.8% |
| *Ischemic* | 22, 64.7% | 18, 52.9% | 33, 63.5% |
| *Other* | 4, 11.7% | 6, 17.6% | 4, 7.6% |
| Hemisphere Affected by Stroke |  |  |  |
| *Left* | 16, 47.1% | 14, 41.2% | 31, 59.6% |
| *Right* | 17, 50.0% | 18, 52.9% | 16, 30.8% |
| *Bilateral* | 1, 2.9% | 0, 0.0% | 3, 5.7% |
| *Unknown* | 0, 0.0% | 2, 5.9% | 2, 3.8% |
| Functional Comorbidity Index | 3.44 (2.05) | 2.97 (1.49) | 3.67 (1.78) |
| Instrumental Activities of Daily Living | 6.82 (1.83) | 7.00 (1.26) | 6.77 (1.78) |
| Mini Mental State Exam | 27.15 (2.56) | 27.56 (2.52) | 27.15 (2.40) |
| Center for Epidemiological Studies-Depression Scale | 9.59 (6.42) | 8.35 (10.30) | 9.92 (7.79) |
| Fugl-Meyer Motor Score | 73.25 (26.48) | 80.09 (24.57) | 87.38 (19.91) |
| Alzheimer's Disease Assessment Scale-Cognitive Plus | 0.39 (0.77) | 0.12 (0.71) | 0.17 (0.88) |
| 13-item Alzheimer's Disease Assessment Scale | 18.16 (7.47) | 16.42 (6.29) | 17.19 (8.00) |
| Motionwatch Sleep Duration (min/night)<sup>1</sup> | 413.46 (60.86) | 434.11 (77.58) | 433.70 (69.68) |
| Motionwatch Sleep Efficiency (%)<sup>1</sup> | 83.29 (8.11) | 84.94 (7.86) | 85.79 (6.69) |
| Pittsburgh Sleep Quality Index | 6.00 (3.09) | 6.38 (3.30) | 5.88 (2.83) |

<sup>1</sup> Means and SDs based on subset of 109 participants (EX=29; ENRICH=31; BAT=49) with Motionwatch measured sleep at baseline

Participant baseline characteristics stratified by device-measured sleep duration and sleep efficiency, and self-reported sleep categorizations are described in Supplementary Materials S2, S3, and S4, respectively. Of the 109 participants with device measured sleep at baseline, we classified 13/29 EX, 17/31 ENRICH, and 28/49 BAT participants as having good device-measured sleep duration at baseline; 15/29 EX, 17/31 ENRICH, and 32/49 BAT participants were classified as having good device-measured efficiency. For self-reported sleep quality, 16/34 EX, 16/34 ENRICH, and 28/52 BAT participants were classified as having good self-reported sleep quality.

### Treatment Moderating Effects of Device-Measured Baseline Sleep Duration

We describe estimated marginal means for each group based on baseline sleep categorization in Table 2. Between-group differences (i.e., EX vs. BAT and ENRICH vs. BAT) are described in Table 3. For participants in either EX or ENRICH with good baseline sleep duration, there were no significant differences in cognitive performance from BAT participants with good baseline sleep duration at either the end of the intervention or at 6-month follow-up. There were no effects of EX or ENRICH on ADAS-Cog-Plus or ADAS-Cog-13 compared with BAT at either the end of the intervention or at 6-month follow-up for participants with poor baseline sleep duration.

**Table 2.**
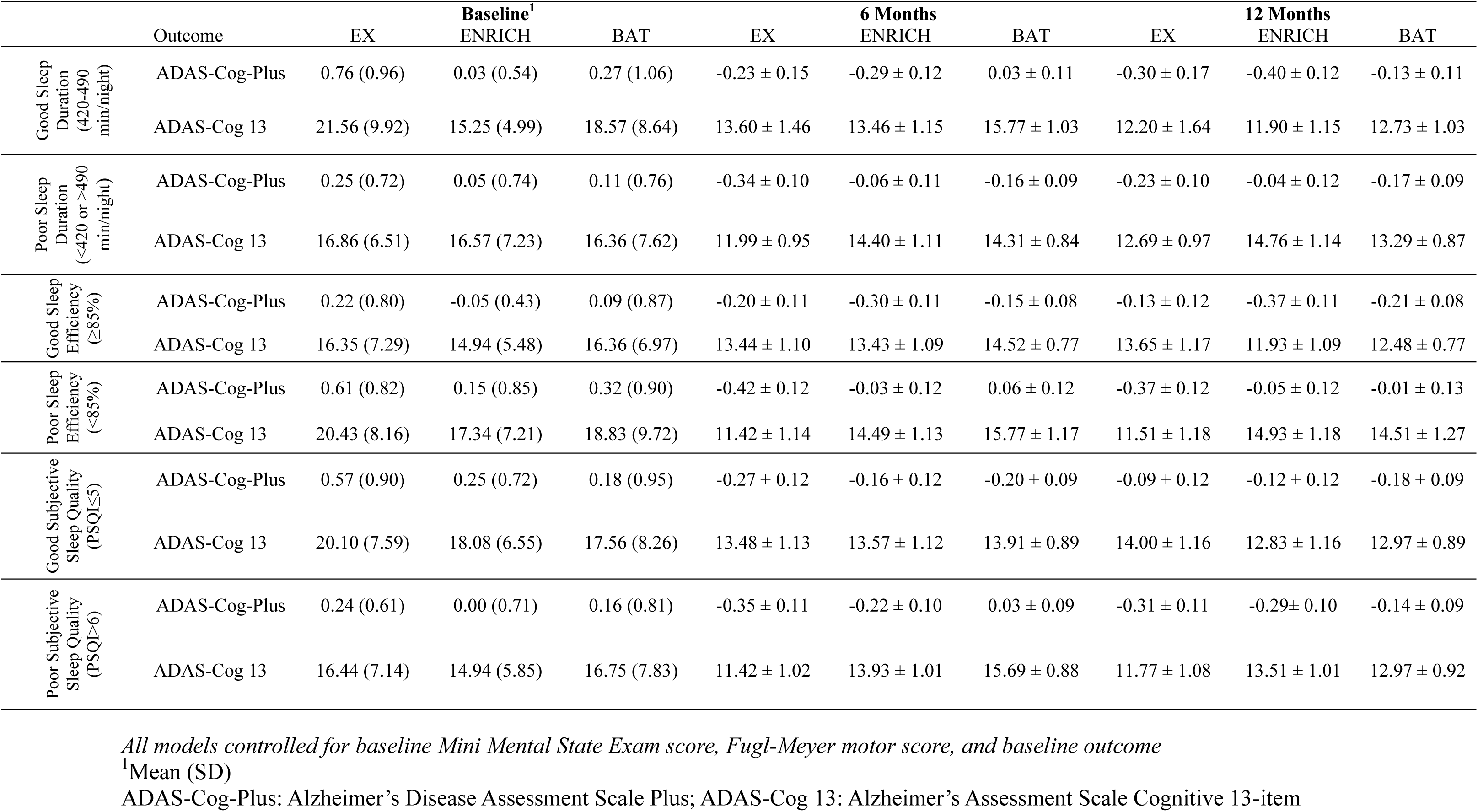
Estimated marginal means ± standard errors for changes in cognitive function and physical function at baseline,6 months (end of intervention) and 12 months (i.e., 6-month follow-up) by treatment group and sleep quality.

|  | Outcome | EX | Baseline<sup>1</sup><br>ENRICH | BAT | EX | 6 Months<br>ENRICH | BAT | EX | 12 Months<br>ENRICH | BAT |
| --- | --- | --- | --- | --- | --- | --- | --- | --- | --- | --- |
| Good Sleep<br>Duration<br>(420-490<br>min/night) | ADAS-Cog-Plus | 0.76 (0.96) | 0.03 (0.54) | 0.27 (1.06) | -0.23 ± 0.15 | -0.29 ± 0.12 | 0.03 ± 0.11 | -0.30 ± 0.17 | -0.40 ± 0.12 | -0.13 ± 0.11 |
| | ADAS-Cog 13 | 21.56 (9.92) | 15.25 (4.99) | 18.57 (8.64) | 13.60 ± 1.46 | 13.46 ± 1.15 | 15.77 ± 1.03 | 12.20 ± 1.64 | 11.90 ± 1.15 | 12.73 ± 1.03 |
| Poor Sleep<br>Duration<br>($<420$ or $>490$<br>min/night) | ADAS-Cog-Plus | 0.25 (0.72) | 0.05 (0.74) | 0.11 (0.76) | -0.34 ± 0.10 | -0.06 ± 0.11 | -0.16 ± 0.09 | -0.23 ± 0.10 | -0.04 ± 0.12 | -0.17 ± 0.09 |
| | ADAS-Cog 13 | 16.86 (6.51) | 16.57 (7.23) | 16.36 (7.62) | 11.99 ± 0.95 | 14.40 ± 1.11 | 14.31 ± 0.84 | 12.69 ± 0.97 | 14.76 ± 1.14 | 13.29 ± 0.87 |
| Good Sleep<br>Efficiency<br>($\geq 85\%$) | ADAS-Cog-Plus | 0.22 (0.80) | -0.05 (0.43) | 0.09 (0.87) | -0.20 ± 0.11 | -0.30 ± 0.11 | -0.15 ± 0.08 | -0.13 ± 0.12 | -0.37 ± 0.11 | -0.21 ± 0.08 |
| | ADAS-Cog 13 | 16.35 (7.29) | 14.94 (5.48) | 16.36 (6.97) | 13.44 ± 1.10 | 13.43 ± 1.09 | 14.52 ± 0.77 | 13.65 ± 1.17 | 11.93 ± 1.09 | 12.48 ± 0.77 |
| Poor Sleep<br>Efficiency<br>($<85\%$) | ADAS-Cog-Plus | 0.61 (0.82) | 0.15 (0.85) | 0.32 (0.90) | -0.42 ± 0.12 | -0.03 ± 0.12 | 0.06 ± 0.12 | -0.37 ± 0.12 | -0.05 ± 0.12 | -0.01 ± 0.13 |
| | ADAS-Cog 13 | 20.43 (8.16) | 17.34 (7.21) | 18.83 (9.72) | 11.42 ± 1.14 | 14.49 ± 1.13 | 15.77 ± 1.17 | 11.51 ± 1.18 | 14.93 ± 1.18 | 14.51 ± 1.27 |
| Good Subjective<br>Sleep Quality<br>(PSQ$\leq 5$) | ADAS-Cog-Plus | 0.57 (0.90) | 0.25 (0.72) | 0.18 (0.95) | -0.27 ± 0.12 | -0.16 ± 0.12 | -0.20 ± 0.09 | -0.09 ± 0.12 | -0.12 ± 0.12 | -0.18 ± 0.09 |
| | ADAS-Cog 13 | 20.10 (7.59) | 18.08 (6.55) | 17.56 (8.26) | 13.48 ± 1.13 | 13.57 ± 1.12 | 13.91 ± 0.89 | 14.00 ± 1.16 | 12.83 ± 1.16 | 12.97 ± 0.89 |
| Poor Subjective<br>Sleep Quality<br>(PSQ$>6$) | ADAS-Cog-Plus | 0.24 (0.61) | 0.00 (0.71) | 0.16 (0.81) | -0.35 ± 0.11 | -0.22 ± 0.10 | 0.03 ± 0.09 | -0.31 ± 0.11 | -0.29 ± 0.10 | -0.14 ± 0.09 |
| | ADAS-Cog 13 | 16.44 (7.14) | 14.94 (5.85) | 16.75 (7.83) | 11.42 ± 1.02 | 13.93 ± 1.01 | 15.69 ± 0.88 | 11.77 ± 1.08 | 13.51 ± 1.01 | 12.97 ± 0.92 |

*All models controlled for baseline Mini Mental State Exam score, Fugl-Meyer motor score, and baseline outcome*
<sup>1</sup>Mean (SD)
ADAS-Cog-Plus: Alzheimer's Disease Assessment Scale Plus; ADAS-Cog 13: Alzheimer's Assessment Scale Cognitive 13-item

**Table 3.** Estimated mean differences and 95% confidence intervals for between-group differences at 6 months (i.e., end of intervention) and 12 months (i.e., 6-month follow-up)

|  | Outcome | Adjusted Between-Group Differences at 6 months (95% CI) |  |  |  | Adjusted Between-Group Differences at 12 months (95% CI) |  |  |  |
| --- | --- | --- | --- | --- | --- | --- | --- | --- | --- |
|  |  | EX vs. BAT | p | ENRICH vs. BAT | p | EX vs. BAT | p | ENRICH vs. BAT | p |
| Good Sleep Duration (420-490 min/night) | ADAS-Cog-Plus | -0.26 (-0.68, 0.03) | 0.286 | -0.32 (-0.68, 0.03) | 0.081 | -0.17 (-0.61, 0.27) | 0.601 | -0.27 (-0.63, 0.08) | 0.156 |
|  | ADAS-Cog 13 | -2.17 (-6.18, 1.84) | 0.378 | -2.31 (-2.67, 3.34) | 0.943 | -0.53 (-4.88, 3.82) | 0.935 | -0.83 (-4.28, 2.62) | 0.799 |
| Poor Sleep Duration (<420 or >490 min/night) | ADAS-Cog-Plus | -0.17 (-0.47, 0.12) | 0.322 | 0.10 (-0.21, 0.42) | 0.676 | -0.07 (-0.37, 0.24) | 0.826 | 0.12 (-0.20, 0.45) | 0.603 |
|  | ADAS-Cog 13 | -2.32 (-5.18, 0.54) | 0.130 | 0.09 (-3.02, 3.19) | 0.994 | -0.60 (-3.54, 2.35) | 0.848 | 1.47 (-1.73, 4.67) | 0.487 |
| Good Sleep Efficiency (≥85%) | ADAS-Cog-Plus | -0.05 (-0.36, 0.25) | 0.877 | -0.15 (-0.46, 0.15) | 0.437 | 0.08 (-0.24, 0.41) | 0.781 | -0.16 (-0.47, 0.14) | 0.393 |
|  | ADAS-Cog 13 | -1.08 (-4.10, 1.94) | 0.631 | -1.10 (-4.09, 1.90) | 0.620 | 1.17 (-1.99, 4.33) | 0.615 | -0.54 (-3.54, 2.45) | 0.873 |
| Poor Sleep Efficiency (<85%) | ADAS-Cog-Plus | -0.48 (-0.85, -0.10) | 0.010 | -0.10 (-0.46, 0.27) | 0.776 | -0.37 (-0.76, 0.02) | 0.069 | -0.05 (-0.43, 0.34) | 0.939 |
|  | ADAS-Cog 13 | -4.34 (-8.00, -0.69) | 0.016 | -1.27 (-4.90, 2.35) | 0.642 | -3.00 (-6.87, 0.87) | 0.153 | 0.42 (-3.43, 4.26) | 0.946 |
| Good Subjective Sleep Quality (PSQI≤5) | ADAS-Cog-Plus | -0.07 (-0.41, 0.27) | 0.835 | 0.04 (-0.30, 0.37) | 0.944 | 0.09 (-0.26, 0.43) | 0.785 | 0.05 (-0.29, 0.39) | 0.904 |
|  | ADAS-Cog 13 | -0.43 (-3.69, 2.82) | 0.924 | -0.34 (-3.59, 2.91) | 0.950 | 1.03 (-2.28, 4.33) | 0.702 | -0.14 (-3.45, 3.17) | 0.989 |
| Poor Subjective Sleep Quality (PSQI>6) | ADAS-Cog-Plus | -0.38 (-0.70, -0.07) | 0.014 | -0.25 (-0.56, 0.06) | 0.129 | -0.18 (-0.51, 0.16) | 0.395 | -0.15 (-0.47, 0.16) | 0.458 |
|  | ADAS-Cog 13 | -4.27 (-7.30, -1.24) | 0.004 | -1.77 (-4.77, 1.24) | 0.323 | -1.20 (-4.38, 1.99) | 0.607 | 0.54 (-2.52, 3.60) | 0.879 |

*All models controlled for baseline Mini Mental State Exam score, Fugl-Meyer motor score, and baseline outcome*
ADAS-Cog-Plus: Alzheimer's Disease Assessment Scale Plus; ADAS-Cog 13: Alzheimer's Assessment Scale Cognitive 13-item

### Treatment Moderating Effects of Device-Measured Baseline Sleep Efficiency

Among participants classified with good baseline sleep efficiency, there were no effects of EX or ENRICH on cognitive performance at either the end of the intervention or at 6-month follow-up. Following the end of the intervention, participants in EX classified with poor baseline sleep efficiency had significantly better ADAS-Cog-Plus (estimated mean difference: −0.48; 95% CI:[−0.85, −0.10]; *p*=0.010) than BAT participants with poor baseline sleep efficiency. There were no between-group differences at 6-month follow-up. We also determined that EX participants with poor baseline sleep efficiency had significantly better ADAS-Cog-13 performance (estimated mean difference: −4.34; 95% CI:[−8.00, −0.69]; *p*=0.016) than BAT participants with poor baseline sleep efficiency following the end of the intervention; there were no between-group differences at 6-month follow-up. There were no effects of ENRICH at either the end of the intervention or 6-month follow-up compared with BAT for participants with poor baseline sleep efficiency.

Post-hoc, we determined that there was a significant difference in the effects of EX vs. BAT on ADAS-Cog-Plus between baseline sleep efficiency categories following the end of the intervention (estimated mean difference: 0.42; 95% CI:[0.01, 0.84]; *p*=0.049), whereby EX participants categorized with poor baseline sleep efficiency had significantly greater improvement in ADAS-Cog-Plus performance than EX with good baseline sleep efficiency. The effects of EX vs. BAT on the ADAS-Cog-13 were not significantly different between baseline sleep efficiency categories at the end of the intervention (estimated mean difference: 3.26; 95% CI:[−0.86, 7.39]; *p*=0.120).

### Treatment Moderating Effects of Baseline Self-Reported Sleep Quality

There were no effects of EX or ENRICH on ADAS-Cog-Plus or ADAS-Cog-13 at either the end of the intervention or at 6-month follow-up for participants with good baseline self-reported sleep quality. Among participants with poor baseline sleep quality, EX had significantly better ADAS-Cog-Plus (estimated mean difference: −0.38; 95% CI:[−0.70, −0.07]; *p=*0.014) and ADAS-Cog-13 performance (estimated mean difference: −4.27; 95% CI:[−7.30, −1.24]; *p*=0.004) than BAT at the end of the intervention. Among participants with poor baseline self-reported sleep quality, there were no effects of ENRICH on cognitive performance compared with BAT at either the end of the intervention or 6-months follow-up. Post-hoc, there were no significant differences between baseline self-reported sleep quality categories in the effects of EX vs. BAT on either ADAS-Cog-Plus (estimated mean difference: 0.31; 95% CI:[−0.09, 0.71]; *p*=0.128) or ADAS-Cog-13 (estimated mean difference: 3.84; 95% CI:[−0.04, 7.12]; *p=*0.052) at the end of the intervention.

## Discussion

We show that adults with chronic stroke and poor sleep may experience greater improvements in cognitive function from EX than their peers with good quality sleep. Sleep does not appear to moderate the effects of ENRICH on cognitive function in adults with chronic stroke.

Among participants classified as having either poor device-measured sleep efficiency (i.e., < 85% efficiency) or poor self-reported sleep quality (PSQI > 6) at baseline, EX had significantly better performance on both the ADAS-Cog-Plus and the ADAS-Cog-13 at trial completion compared with those in BAT. Importantly, EX with poor baseline sleep efficiency improved on the ADAS-Cog-13 by 4.34 points compared with BAT with poor baseline sleep efficiency; EX with poor baseline self-reported sleep quality improved by 4.27 points compared with BAT with poor baseline self-reported sleep quality. Each of these improvements exceed the established minimally clinically important difference (i.e., ≥3.0 points).^30^ Post-hoc comparisons indicated EX categorized with poor baseline sleep efficiency had a significantly greater improvement in ADAS-Cog-Plus performance than EX with good baseline sleep efficiency. Neither good (i.e., 420-490 minutes/night) nor poor baseline sleep duration (<420 minutes/night or >490 minutes/night) moderated the effects of EX on cognitive function.

For ENRICH, baseline sleep did not moderate the effects of the intervention on cognitive function. In the primary analysis of this study, ENRICH did not improve cognitive function.^4^

Our results highlight that baseline sleep quality, but not duration, moderates the effects of EX on cognitive function in older adults with chronic stroke. Bloomberg and colleagues^35^ determined that greater amounts of physical activity does not appear to ameliorate the negative consequences of short sleep duration on cognitive function. It is unclear how longer sleep durations (i.e., >8 hours) impact the effects of EX on cognitive function. By comparison, Lambaise and colleagues^36^ found that greater physical activity attenuated the negative impacts of poor sleep on cognitive function. Hence, we suggest that the effects of EX on cognitive function in people with chronic stroke may be most potent among individuals who are suffering from poor sleep quality.

### Limitations

This was a secondary analysis wherein we stratified our results based on sleep categorization. We did not adjust for multiple comparisons due to the exploratory nature of this study and our stratified treatment groups were not fully powered. Our sample had heterogeneous stroke types and locations, which may be linked to different sleep disturbances and cognitive issues.^37^ ^38^ We did not take take steps to ensure diversity in the sample or the trial steering committee.

There is not yet criterion evidence of validity for the MW8 among adults with chronic stroke; in a previous investigation,^14^ we determined that MW8 provides reliable estimates of sleep duration and efficiency among older adults with and without cognitive impairment. We queried whether participants were diagnosed with OSA, but it is still plausible that some participants had undiagnosed OSA. Other sleep disorders which we did not query about (e.g., restless leg syndrome) could have also confounded our results. The study sample only included chronic stroke survivors with mild to moderate motor impairments. Finally, due to the diverse content of the ENRICH intervention, there may have been insufficient dose and specificity of training to elicit an effect.

### Conclusions

The findings from this secondary analysis of a RCT suggest that baseline sleep quality moderates the effects of EX on cognitive function in people with chronic stroke, and can induce clinically important improvements in cognitive performance in this clinical population at risk for dementia. Adults with chronic stroke and poor sleep may thus be a key target population for promoting cognitive health through exercise training.

## Data Availability

De-identified participant data will be made available by the corresponding author to others who propose a reasonable scientific request and obtain appropriate ethics.

